# Racial and Ethnic Disparities in Healthcare Utilization and Mortality by Neighborhood Poverty among Individuals with Congenital Heart Defects, four U.S. Surveillance Sites, 2011-2013

**DOI:** 10.1101/2023.12.12.23299887

**Authors:** Cheryl L. Raskind-Hood, Vijaya Kancherla, Lindsey C. Ivey, Fred H. Rodriguez, Anaclare M. Sullivan, George K. Lui, Lorenzo Botto, Marcia Feldkamp, Jennifer S. Li, Alfred D’Ottavio, Sherry L. Farr, Jill Glidewell, Wendy M. Book

## Abstract

**BACKGROUND:** Socioeconomic factors may lead to a disproportionate impact on healthcare utilization and mortality among individuals with congenital heart defects (CHD) by race, ethnicity, and socio-economic factors. How neighborhood poverty affects racial and ethnic disparities in healthcare utilization and mortality among individuals with CHD across the lifespan is not well described.

**METHODS:** Individuals, 1-64 years, with at least one CHD-related ICD-9-CM code were identified from healthcare encounters between 01/01/2011-12/31/2013 from four U.S. sites. Residence was classified into lower or higher poverty neighborhoods based on ZCTA from the 2014 American Community Survey 5-Year Estimates. Multivariable logistic regression models, adjusting for site, sex, CHD anatomic severity, and insurance evaluated associations between race and ethnicity, and healthcare utilization and mortality, stratified by neighborhood poverty.

**RESULTS:** Of 31,542 individuals, 22.2% were non-Hispanic Black (nHB) and 17.0% Hispanic. In high poverty neighborhoods, nHB (44.4%) and Hispanic (47.7%) individuals, respectively, were more likely to be hospitalized (aOR)=1.2 [95%CI=1.0-1.3] and aOR=1.3 [95%CI=1.2-1.5]) and have ED visits (aOR=1.3 [95%CI=1.2-1.5] and aOR=1.7 [95%CI=1.5-2.0]) compared to non-Hispanic White (nHW) individuals. In high poverty neighborhoods, nHB individuals with CHD had 1.7 times the odds of mortality compared to nHW individuals in high poverty neighborhoods (95%CI=1.1-2.7). Racial and ethnic disparities in healthcare utilization were similar in low poverty neighborhoods, but disparities in mortality were attenuated (aOR for nHB=1.2 [95%CI=0.9-1.7]).

**CONCLUSIONS:** Racial and ethnic disparities in healthcare utilization were found among individuals with CHD in low and high poverty neighborhoods, but mortality disparities were larger in high poverty neighborhoods. Understanding individual- and community-level social determinants of health, including access to healthcare, may help address racial and ethnic inequities in healthcare utilization and mortality among individuals with CHD.

## INTRODUCTION

Congenital heart defects (CHD) affect approximately 1% of all births in the U.S.^1^ Advances in surgery, technology, and peri-operative care for children with CHD have improved survival to adulthood,^2, 3, 4, 5, 6, 7, 8^ resulting in more adults than children with CHD in the U.S.^9, 10^ Between 1998 and 2005, the number of hospital admissions for adults with CHD more than doubled^11^ and mortality from CHD has decreased,^12^ yet racial and ethnic and socioeconomic disparities in healthcare utilization and mortality exist.

Social determinants of health are associated with racial and ethnic disparities in healthcare utilization and mortality in the population, and disparities in outcomes have been described among children with CHD. Children with CHD within racial and ethnic minority groups, compared to White children with CHD, have been found to experience greater healthcare lapses among those who underwent CHD surgery,^13^ higher severity of illness scores,^14^ increased odds of complications,^15, 16^ greater odds of surgery at lower volume CHD surgical centers for those with hypoplastic left heart syndrome^8^, and higher mortality.^14, 15, 17, 18, 19^ The National Institute of Minority Health and Health Disparities Research Framework considers multiple domains and levels of influence to conceptualize health disparities, including the role of community environment and resources, and health care policies to better understand the relationship of structural discrimination and health disparities.^20, 21^

Socioeconomic factors and neighborhood poverty are key social determinants of health associated with healthcare utilization and mortality among individuals with CHD. While previous studies have shown a decrease in mortality from CHD,^12, 22, 23^ studies have shown disparities in mortality by race, ethnicity, and type of health insurance.^12, 24^ Among children undergoing CHD surgery, those with private insurance had improved outcomes,^12, 15, 16, 25, 26^ and those from low, compared to high, income neighborhoods had higher mortality.^27^ Similarly, among adolescents and adults with CHD in a Colorado cohort,^28^ poverty was associated with higher rates of hospitalization, emergency department (ED) visits, and adverse cardiac outcomes.

Few studies have examined how social determinants of health, including neighborhood poverty, mediate the relationship between race and ethnicity and healthcare utilization and mortality among individuals, including adults, with CHD. One study among CHD individuals examined the modifying effect of neighborhood household income on the relationship between race and mortality, finding that Black individuals had longer length of stay and higher mortality compared to their White counterparts, with mortality potentiated by lower neighborhood income.^29^ A review by Richardson et al. (2021) showed significant racial and ethnic disparity in healthcare utilization among individuals with CHD, and socioeconomic factors mediated the risk.^15^ To expand knowledge on this topic, this study aims to examine the association between race and ethnicity and healthcare utilization (outpatient visits, hospitalizations, and ED visits) and mortality, by neighborhood poverty status, among individuals with CHD, aged 1-64 years. Study findings may help determine disparities in morbidity and mortality by race and ethnicity and neighborhood poverty status among people with CHD.

## METHODS

This retrospective study of children and adults with CHD used data from four sites (Georgia [GA], North Carolina [NC], New York [NY], and Utah [UT]) participating in a collaborative CHD surveillance project funded by the U.S. Centers for Disease Control and Prevention (CDC), CDC-RFA-DD15-1506. Individuals with CHD were identified based on at least one CHD-related *International Classification of Disease version 9.0 Clinical Modification* (ICD-9-CM) code (Appendix A), within the 745.xx-747.xx range. These codes were identified from electronic administrative and clinical sources, state Medicaid claims, state vital records and birth defects registries. Compilation and sharing of de-identified data with the CDC were approved by each site’s Institutional Review Board. A detailed methodology of the parent project has been published.^30^

### CHD case classification

CHD diagnostic codes, which are based on native CHD anatomy, were categorized into one of five mutually exclusive hierarchical CHD severity groupings similar to the Marelli et al. (2007) classification scheme,^2^ integrating both hemodynamic severity and basic anatomy: 1) severe; 2) shunt (excluding isolated 745.5 secundum atrial septal defect/patent foramen ovale); 3) valve; 4) shunt and valve lesions; and 5) ‘Other’ CHDs (**Appendix A**). Severe CHD includes endocardial cushion defects (745.6 / 745.60 / 745.69), interrupted aortic arch (747.11), tetralogy of Fallot (745.2), total anomalous pulmonary venous return (747.41), tricuspid atresia (746.1), transposition complexes (745.1 / 745.10 / 745.11 / 745.12 / 745.19), truncus arteriosus (745.0) and univentricular hearts (745.3). Individuals with multiple CHD-related ICD-9-CM codes who had at least one severe code were classified as having a severe condition regardless of the number of non-severe codes they had.

### Inclusion and Exclusion Criteria

The 2011-2013 population included 72,433 individuals with one or more healthcare encounters during the surveillance period with a CHD-related ICD-9-CM code. Exclusions included those: 1) whose age was < 1 year or greater than 64 years (n=10,181 excluded, where (10,181/72,443=14.1% of total leaving 62,252; 2) diagnosed with a 745.5 code in isolation (secundum atrial septal defect/patent foramen ovale) or in combination with either 746.89 (Other specified anomalies of heart: Other) or 746.9 (Unspecified anomaly of heart) (n=14,228 excluded, where 14,228/62,252=22.8% of remaining total leaving 48,024) or any “other CHD” code (n=504 excluded, where 504/48,024=1.0% of remaining total leaving 47,520); 4) diagnosed with only “other CHD” codes (n=11,381 excluded, where 11,381/47,520=23.9% of remaining total leaving 36,139), as these codes are known to have poor positive predictive value for CHD;^31, 32^ 5) with unknown sex (n=1 excluded, where 1/36,139 = <1.0% of remaining total leaving 36,138); 6) with unknown neighborhood income status (n=249 excluded, where 249/36,138=0.7% of remaining total leaving 35,889); 7) with unknown neighborhood poverty status (n=1 excluded, where 1/35,889=<1.0% of remaining total leaving 35,888); and 8) with unknown race and ethnicity (n=4,346 excluded, where 4,346/35,888=12.1% of remaining total leaving 31,542).

After implementing above exclusions, 31,542 individuals with probable CHD aged 1-64 years, who: 1) had at least one healthcare encounter between 1/1/2011-12/31/2013; 2) had at least one ICD-9-CM CHD-related code documented (see **Appendix A**); and 3) resided in the four site-specific catchment areas were eligible and included in the analysis. Site-specific catchment areas spanned the entire state for individuals that resided in NC and UT. For GA, data were collected from individuals residing within one of the five metropolitan-Atlanta counties (Clayton, Cobb, DeKalb, Fulton, and Gwinnett), and for NY, individuals resided in one of 11 NY counties (Allegany, Bronx, Cattaraugus, Chautauqua, Erie, Genesee, Monroe, Niagara, Orleans, Wyoming, and Westchester).

### Study Variables

#### Healthcare utilization (Outcome Variable)

Healthcare utilization was categorized as outpatient visits, hospitalizations, and ED visits. If an ED visit led to a hospitalization, the encounter was captured as a hospitalization. Separately by encounter type, the number of encounters were calculated by combining overlapping date ranges into a single encounter, summing encounters over the surveillance period, and dichotomizing as 0 and ≥1. When dates of different encounter types overlapped, a hierarchical scheme was applied in the following order: 1) inpatient hospitalizations; 2) ED visits; and 3) outpatient visits.

#### Mortality (Outcome Variable)

Mortality status was determined by deterministically linking individuals to state-specific death certificates at each site. If the individual did not match to a death record during the surveillance period, the person was considered alive.

#### Race and Ethnicity

Race and ethnicity were based on data recorded in the electronic health records (eHR) and categorized as non-Hispanic White (nHW), non-Hispanic Black (nHB), Hispanic, and other race [Asian, American Indian/Native American, Native Hawaiian/Pacific Islander, and multi-racial (excluding Black multi-racial)]. Individuals could have more than one race recorded. For this analysis, individuals with nHB race were categorized as nHB, even if they had other races recorded. All other individuals with multiple races recorded were categorized as multi-racial. (n=161/31,542). For purposes of analysis, racial groups comprising a small percentage of the dataset were combined into a category called “Other” because the sample sizes were too small.

#### Neighborhood Economic Status

Two metrics were used to examine neighborhood economic status: 1) neighborhood median income; and 2) neighborhood poverty status. Values for each metric were based on individuals’ ZIP Code Tabulation Area (ZCTA) from the 2014 American Community Survey (ACS) 5-year estimates, estimated across years 2010-2014.^33^ Annual neighborhood median income in USD (U.S. dollar currency) was classified into 3 groups: <$40,000, $40,000-$75,000, and >$75,000. Neighborhood poverty status was defined as the percent of households in the ZCTA below 100% of the federal poverty level (FPL) and was classified into low neighborhood poverty (≤25% of households below FPL) and high neighborhood poverty (>25% of households below FPL).

#### Other Co-variables

Individual-level covariates were determined based on literature review and included site, age, sex, CHD anatomic severity, and health insurance. Age in years was calculated by subtracting the individual’s date of birth from the date of the individual’s first healthcare encounter during the 2011-2013 surveillance period where an eligible CHD-related ICD-9-CM code appeared. Age was categorized into four groups: 1-10 years, 11-18 years, 19-44 years, and 45-64 years. Sex was classified as male or female. CHD anatomic severity was categorized as: 1) severe; 2) shunt (excluding isolated 745.5 secundum atrial septal defect/patent foramen ovale); 3) valve; and 4) shunt and valve lesions. Health insurance status was classified as: 1) “any public”, if health insurance was documented at any healthcare encounter as Medicaid or Medicare; 2) “private”, if health insurance was documented at all healthcare encounters as a private company, other government which includes military, Veterans Affairs, Tricare, and other federal employee insurance benefits or other health insurance coverage; 3) “none”, if all healthcare encounters indicated self-pay, uninsured or “no insurance coverage”; and 4) “unknown”, when health insurance status was unavailable.

### Statistical Analysis

SAS version 9.4 (SAS Institute Inc., Cary, NC) was used for all analyses. Descriptive analyses were conducted to examine the frequency distributions and percentages of individual characteristics by race and ethnicity. Bivariate analyses were conducted to describe and compare the associations between race and ethnicity, the covariables, and healthcare utilization (inpatient hospitalizations, ED visits, and outpatient visits) and mortality, using two-sided Chi-square tests. *P* values at <0.05 were considered statistically significant. Effect modification was considered a priori by neighborhood poverty status. Adjusted odds ratios (aORs) and 95% confidence intervals (95% CIs) were estimated using multivariable logistic regression analysis. Potential confounders were selected applying a 10% change-in-estimate criterion. Models were stratified by neighborhood poverty status.

## RESULTS

There were 31,542 individuals with CHD identified from four surveillance sites who were eligible for analysis. Overall, 57.6% were nHW, 22.2% were nHB, 17.0% were Hispanic, and 3.2% were another race and ethnicity (**Table 1**). Distribution of all demographics and healthcare types (p<0.0001) and mortality (p=0.0007) differed by race and ethnicity. Among nHW individuals with CHD, 32.0% were 1-10 years of age as compared to 51.3% among nHB and 51.5% among Hispanic individuals with CHD. The proportion of CHD individuals ages 45-64 years was lower than all other age groups in all racial and ethnic groups. Among nHW individuals with CHD, 18.9% were 45-64 years of age as compared to 10.9% and 10.5% for nHB and Hispanic individuals, respectively. Overall median age was 14.0 years. Among nHW individuals with CHD, valve lesions were the most common (44.8%), while shunt lesions were most prevalent among nHB (35.1%), and Hispanic (38.7%) individuals. Additionally, 33.9% of nHW individuals with CHD had public health insurance compared to 71.3% of nHB and 82.0% of Hispanic individuals. Similar patterns of neighborhood median income and poverty status emerged, with less than 20% of nHW and close to half of nHB and Hispanic individuals living in lower income/higher poverty neighborhoods. For all characteristics, values for individuals with “other” race and ethnicity generally mirrored those of nHB and Hispanic individuals or fell between those of nHW and NHB/Hispanic individuals. Among individuals with CHD, 93.1% had at least one outpatient visit, 42.9% had at least one inpatient hospitalization, and 34.2% visited the ED at least once (**Table 1**). All three encounter types differed significantly by race and ethnicity (p<0.0001 for all). Outpatient care was the most frequent across all racial and ethnic groups. Overall, mortality of individuals with CHD during the surveillance period was 1.2%, ranging from 0.9% for Hispanic individuals to 1.6% for nHB (p=0.0007) (**Table 1**). Patterns in mortality among nHW, nHB, and Hispanic individuals were similar for younger (1-18 years; nHW: 0.5%; nHB: 0.8%, Hispanic: 0.6%; p<0.05) and older age groups (19-64 years; nHW: 1.8%; nHB: 3.5%, Hispanic: 1.7%; p<0.0001), with nHB individuals having higher mortality rates than nHW and Hispanic individuals (data not shown).

**Table 1.** Characteristics of individuals with congenital heart defects, total and by racial and ethnic group.

|  | Total<br>N=31542 | Non-Hispanic<br>White<br>n=18173<br>(57.6%) | Non-Hispanic<br>Black<br>n=6991<br>(22.2%) | Hispanic<br>n=5365<br>(17.0%) | Other<sup>†</sup><br>n=1013<br>(3.2%) | P-value<sup>§</sup> |
| --- | --- | --- | --- | --- | --- | --- |
| Characteristics | n (%) | n (%)<sup>*</sup> | n (%)<sup>*</sup> | n (%)<sup>*</sup> | n (%)<sup>*</sup> |  |
| Site |  |  |  |  |  |  |
| Georgia | 6070 (19.2) | 2854 (15.7) | 2415 (34.5) | 522 (9.7) | 279 (27.5) | <0.0001 |
| North Carolina | 11830 (37.5) | 8033 (44.2) | 2454 (35.1) | 981 (18.3) | 362 (35.7) |  |
| New York | 10240 (32.5) | 4247 (23.4) | 2091 (29.9) | 3534 (65.9) | 368 (36.3) |  |
| Utah | 3402 (10.8) | 3039 (16.7) | 31 (0.4) | 328 (6.1) | <10 (--) |  |
| Age<sup>†</sup> (years) |  |  |  |  |  |  |
| 1-10 | 12700 (40.3) | 5815 (32.0) | 3585 (51.3) | 2765 (51.5) | 535 (52.8) | <0.0001 |
| 11-18 | 6251 (19.8) | 3842 (21.2) | 1311 (18.7) | 943 (17.6) | 155 (15.3) |  |
| 19-44 | 7705 (24.4) | 5076 (27.9) | 1333 (19.1) | 1096 (20.4) | 200 (19.8) |  |
| 45-64 | 4886 (15.5) | 3440 (18.9) | 762 (10.9) | 561 (10.5) | 123 (12.1) |  |
| Sex |  |  |  |  |  |  |
| Female | 15376 (48.8) | 8295 (46.6) | 3718 (53.2) | 2834 (52.8) | 529 (52.2) | <0.0001 |
| Male | 16166 (51.2) | 9878 (54.4) | 3273 (46.8) | 2531 (47.2) | 484 (47.8) |  |
| CHD Anatomic Severity |  |  |  |  |  |  |
| Severe | 7815 (24.8) | 4488 (24.7) | 1829 (26.2) | 1230 (22.9) | 268 (26.5) | <0.0001 |
| Shunt | 9067 (28.8) | 4163 (22.9) | 2452 (35.1) | 2076 (38.7) | 376 (37.1) |  |
| Valve | 12059 (38.2) | 8145 (44.8) | 2052 (29.3) | 1601 (29.8) | 261 (25.8) |  |
| Shunt and Valve | 2601 (8.2) | 1377 (7.6) | 658 (9.4) | 458 (8.6) | 108 (10.6) |  |
| Health Insurance |  |  |  |  |  |  |
| Any public | 16071 (51.0) | 6157 (33.9) | 4987 (71.3) | 4399 (82.0) | 528 (52.1) | <0.0001 |
| Private | 12561 (39.8) | 10063 (55.4) | 1440 (20.6) | 700 (13.1) | 358 (35.3) |  |
| None | 310 (1.0) | 148 (0.8) | 87 (1.2) | 67 (1.3) | <10 (--) |  |
| Unknown | 2600 (8.2) | 1805 (9.9) | 477 (6.8) | 199 (3.7) | 119 (11.8) |  |

| **Neighborhood median income (USD)** |  |  |  |  |  |  |
| --- | --- | --- | --- | --- | --- | --- |
| <\$40,000 | 10003 (31.7) | 3295 (18.1) | 3313 (47.4) | 3039 (56.7) | 356 (35.1) | <0.0001 |
| \$40,000-\$75,000 | 17031 (54.0) | 11142 (61.3) | 3398 (48.6) | 2035 (37.9) | 456 (45.0) | |
| >\$75,000 | 4508 (14.3) | 3736 (20.6) | 280 (4.0) | 291 (5.4) | 201 (19.9) | |
| **Neighborhood poverty status** |  |  |  |  |  |  |
| Low poverty neighborhood<br>(<25% households below FPL) | 21234 (67.3) | 14684 (80.8) | 3587 (51.3) | 2328 (43.4) | 635 (62.7) | <0.0001 |
| High poverty neighborhood<br>(≥25% households below FPL) | 10308 (32.7) | 3489 (19.2) | 3404 (48.7) | 3037 (56.6) | 378 (37.3) |  |
| **Healthcare utilization** |  |  |  |  |  |  |
| Outpatient visits | 29352 (93.1) | 16730 (92.1) | 6532 (93.4) | 5175 (96.5) | 915 (90.3) | <0.0001 |
| Hospitalizations | 13523 (42.9) | 7318 (40.3) | 3099 (44.3) | 2599 (48.4) | 507 (50.1) | <0.0001 |
| Emergency Department visits | 10778 (34.2) | 4827 (26.6) | 2453 (35.1) | 3226 (60.1) | 272 (26.9) | <0.0001 |
| **Mortality** |  |  |  |  |  |  |
| Yes | 374 (1.2) | 197 (1.1) | 115 (1.6) | 50 (0.9) | 12 (1.2) | 0.0007 |

**Acronyms:** CHD = congenital heart defects; FPL = Federal Poverty Level; USD = United States dollar currency.
\* Column percentages reported in cells.
† Estimates for 1-10-year-olds based on three sites: five counties in metropolitan-Atlanta, GA; 11 counties in NY state; and statewide in NC; estimates for 11-18, 19-44, and 45-64-year-olds based on four sites: five counties in metropolitan-Atlanta, GA; 11 counties in NY; and statewide in NC and UT.
‡ Other racial and ethnic group includes Asian, American Indian/Native American, Native Hawaiian or Other Pacific Islander, and multi-racial.
§ Comparing racial and ethnic groups only; analysis does not include “Total” column.
|| Insurance categorization: public defined as Medicaid or Medicare; private defined as private, other government, or other insurance; none defined as self-pay/uninsured; and unknown defined as unavailable, unknown, or no insurance indicated.
Note. Overall, median age was 14.0 years.

**Table 2** presents stratified results by neighborhood poverty status in an effort to examine effect modification. Within high poverty neighborhoods, nHB and Hispanic individuals together (62.5%; 33.0% nHB and 29.5% Hispanic) accounted for the largest proportion of individuals compared to nHW individuals (33.9%) or other races (3.7%), whereas in low poverty neighborhoods, nHW (69.2%) comprised the majority of individuals compared to the 27.9% of nHB and Hispanic individuals combined (16.9% nHB and 11.0% Hispanic) or other races (3.0%), p<0.0001) (data not shown). Among those in high poverty neighborhoods, nHB (44.4%), Hispanic individuals (47.7%), and individuals of other race and ethnicity (55.0%), respectively, had 1.2 (95% CI=1.0-1.3), 1.3 (95% CI 1.2-1.5) and 1.7 (95% CI 1.3-2.1) times higher odds of being hospitalized than nHW (42.3%; referent), respectively, after adjusting for confounders. Non-Hispanic Black [38.9%; aOR=1.3 (95% CI=1.2-1.5)] and Hispanic individuals [66.0%; aOR=1.7 (95% CI=1.5-2.0)] also had higher adjusted odds of ED visits compared to nHW individuals (26.2%; referent). In addition, in high poverty neighborhoods, 1.1% of nHW (referent), 1.9% of nHB [aOR=1.7 (95% CI=1.1-2.7)], and 0.9% of Hispanic individuals [aOR=0.7 (95% CI=0.4-1.3)] died during the surveillance period. Among those in low poverty neighborhoods, associations between race and ethnicity and healthcare utilization were of similar magnitude and direction as those observed in high poverty neighborhoods, with the exceptions that Hispanic individuals (95.8%) had 1.4 (95% CI=1.1-1.7) times higher adjusted odds of outpatient visits compared to nHW (92.0%; referent), and associations between mortality and race and ethnicity between nHB (1.5%) and nHW (1.1%) were attenuated [nHB: aOR=1.2 (95%CI=0.9-1.7)].

**Table 2.** Associations between race and ethnicity and healthcare utilization and mortality, by neighborhood poverty status among individuals with congenital heart defects.

| **High poverty neighborhood**<br>**(≥25% households below Federal Poverty Level\*)** |  |  |  |  |  |  |  |  |
| --- | --- | --- | --- | --- | --- | --- | --- | --- |
|  | **Outpatient visits\*\*** |  | **Hospitalizations\*\*** |  | **Emergency Department visits\*** |  | **Mortality** |  |
|  | **n**<br>**(%)** | **aOR<sup>†</sup>**<br>**(95% CI)** | **n**<br>**(%)** | **aOR<sup>†</sup>**<br>**(95% CI)** | **n**<br>**(%)** | **aOR<sup>†</sup>**<br>**(95% CI)** | **n**<br>**(%)** | **aOR<sup>†</sup>**<br>**(95% CI)** |
| Non-Hispanic White | 3221<br>(92.3) | 1.0<br>(referent) | 1475<br>(42.3) | 1.0<br>(referent) | 914<br>(26.2) | 1.0<br>(referent) | 38<br>(1.1) | 1.0<br>(referent) |
| Non-Hispanic Black | 3207<br>(94.2) | 0.9<br>(0.7-1.1) | 1511<br>(44.4) | 1.2<br>(1.0-1.3) | 1325<br>(38.9) | 1.3<br>(1.2-1.5) | 63<br>(1.9) | 1.7<br>(1.1-2.7) |
| Hispanic | 2944<br>(96.9) | 1.0<br>(0.7-1.3) | 1449<br>(47.7) | 1.3<br>(1.2-1.5) | 2004<br>(66.0) | 1.7<br>(1.5-2.0) | 27<br>(0.9) | 0.7<br>(0.4-1.3) |
| Other<sup>‡</sup> | 342<br>(90.5) | 0.6<br>(0.4-0.9) | 208<br>(55.0) | 1.7<br>(1.3-2.1) | 119<br>(31.5) | 0.7<br>(0.5-0.9) | <10<br>-- | 1.0<br>(0.3-2.8) |
| **Low poverty neighborhood**<br>**(<25% households below Federal Poverty Level\*)** |  |  |  |  |  |  |  |  |
|  | **Outpatient visits\*\*** |  | **Hospitalizations\*\*** |  | **Emergency Department visits\*\*** |  | **Mortality** |  |
|  | **n**<br>**(%)** | **aOR<sup>†</sup>**<br>**(95% CI)** | **n**<br>**(%)** | **aOR<sup>†</sup>**<br>**(95% CI)** | **n**<br>**(%)** | **aOR<sup>†</sup>**<br>**(95% CI)** | **n**<br>**(%)** | **aOR<sup>†</sup>**<br>**(95% CI)** |
| Non-Hispanic White | 13509<br>(92.0) | 1.0<br>(referent) | 5843<br>(39.8) | 1.0<br>(referent) | 3913<br>(26.7) | 1.0<br>(referent) | 159<br>(1.1) | 1.0<br>(referent) |
| Non-Hispanic Black | 3325<br>(92.7) | 0.9<br>(0.8-1.0) | 1588<br>(44.3) | 1.2<br>(1.1-1.3) | 1128<br>(31.5) | 1.6<br>(1.5-1.8) | 52<br>(1.5) | 1.2<br>(0.9-1.7) |
| Hispanic | 2231<br>(95.8) | 1.4<br>(1.1-1.7) | 1150<br>(49.4) | 1.5<br>(1.3-1.6) | 1222<br>(52.5) | 2.0<br>(1.8-2.2) | 23<br>(1.0) | 0.7<br>(0.5-1.2) |
| Other<sup>‡</sup> | 573<br>(90.2) | 0.9<br>(0.7-1.2) | 299<br>(47.1) | 1.4<br>(1.2-1.7) | 153<br>(24.1) | 1.0<br>(0.8-1.2) | <10<br>-- | 1.4<br>(0.7-2.9) |

**Acronyms:** aOR = adjusted odds ratio; CI = confidence interval; CHD = congenital heart defect
\*Assessed based on individuals' ZIP Code Tabulation Area (ZCTA) from the 2014 American Community Survey (ACS) 5-year estimates, estimated across years 2010-2014 (U.S. Census Bureau, 2014).
\*\* Zero counts for visit types are not included; healthcare utilization visit categories are not mutually exclusive.
<sup>†</sup> Multivariate analyses adjusted for age, sex, insurance, 4-level CHD severity group, and surveillance site.
<sup>‡</sup> Other racial and ethnic group includes Asian, American Indian/Native American, Native Hawaiian or Other Pacific Islander, and multi-racial.

## DISCUSSION

In this 3-year multi-state health administrative data-based study of race and ethnic disparities in healthcare utilization and mortality among children and adults with CHD aged 1-64 years, we found that nHB and Hispanic individuals were significantly more likely to have hospitalizations and have ED visits compared to nHW individuals, irrespective of their neighborhood poverty status. However, when examining mortality, nHB individuals in high poverty neighborhoods had a nearly 2 times higher odds of death compared to nHW, but this disparity in mortality was attenuated for those living in low poverty neighborhoods. This study not only adds to the current understanding of the role of neighborhood poverty contributing towards racial and ethnic disparities in healthcare utilization among individuals with CHD in the U.S. but also extends the understanding of how neighborhood poverty status modifies those associations. Findings from the study are representative of the populations that use healthcare services living in regions with similar socioeconomic and demographic profiles.

We did not find any previous studies that examined the association between race and ethnicity and healthcare utilization, or mortality, for adults up to age 64 years with CHD, stratified by neighborhood poverty status. In a study using data from the Pediatric Health Information System among individuals <26 years, Karamlou et al. (2022) found the association between race and ethnicity and mortality to be significant among those with low median neighborhood income (a measure often considered equivalent to neighborhoods of higher poverty),^29^ similar to our finding showing a significant association between race and ethnicity and mortality among people with CHD residing in high poverty neighborhoods. In our study among individuals aged 1-64 years, the racial and ethnic disparity in mortality was attenuated for those living in low poverty neighborhoods. While findings are similar, the studies are not directly comparable due to differences in the age of individuals in the two studies, where the prior study limited their individuals to age <26 years,^29^ while our study sample consisted of individuals aged 1-64 years.

Similar to findings from other studies,^12, 34^ mortality was lower for Hispanic individuals compared to nHW individuals and higher for nHB individuals compared to nHW individuals in our study. Our study was one of the first to adjust for CHD anatomic severity along with age, sex, surveillance site, and insurance status, and stratify by neighborhood poverty status. Our analysis did not look at mortality prior to 1 year of age. Karamlou et al. (2022) showed increased mortality among nHB neonates compared to neonates of other racial groups including nHW, Hispanic, Asian, American Indian, and Pacific Islander individuals, and the association was modified by neighborhood household income.^29^ Our study found that racial and ethnic differences in mortality for people with CHD persist beyond the first year of life, particularly in higher poverty neighborhoods.

Our study found that in high poverty neighborhoods, both nHB and Hispanic individuals with CHD experienced higher ED use compared to nHW individuals. In a recent study by Benavidez et al. (2019), individuals of Hispanic ethnicity and CHD were significantly more likely to experience hospital re-admissions compared to individuals who do not identify as Hispanic.^35^ Post-operative mortality by race was not predicted by accessing care alone. Barriers impacting healthcare access or long gaps in care are often associated with complications for selected types of CHDs.^7, 36^ While rates of health insurance have improved for adults with CHD since the Affordable Care Act (ACA) was implemented,^37^ gaps in care utilization persist, with about 50% of adolescents with CHD experiencing barriers to transitioning from pediatric to adult CHD specialty care.^5^

While gaps in CHD specialty care have been associated with adverse outcomes,^5, 38^ our data show increased hospitalizations and ED visits among nHB and Hispanic individuals in both low and high poverty neighborhoods. In addition, no difference by race and ethnicity in outpatient visits in high poverty neighborhoods were revealed, while in low poverty neighborhoods, Hispanic individuals had higher outpatient utilization. We were not able to determine the provider specialty for outpatient visits; thus, while there was little difference in having one or more outpatient encounters by race and ethnicity, there may be differences in the number of outpatient visits and/or in the number of congenital cardiology outpatient encounters. It is possible that individuals with complications due to CHD may present to the ED in a more advanced disease state that could contribute to increased mortality. Surgical complications are an additional factor associated with high healthcare utilization in terms of hospital readmissions; efforts to reduce such complications have been proposed to decrease healthcare utilization and associated costs.^39, 40^

Ongoing regular care with a congenital cardiologist reduces mortality^2, 38^ and improves outcomes across the CHD anatomic severity levels.^41^ Individuals may be out of routine congenital cardiology care for a variety of socioeconomic reasons, including limited access to transportation to centralized tertiary care centers, lack of insurance access, language barriers, lack of paid sick leave, inability to take time off work, economic constraints and limited understanding of the purpose and benefits of routine surveillance care for their CHD.^5^ Thus, individuals who are out of specialty care may seek care through the ED at a higher rate and in a sicker state than those individuals who have remained in congenital cardiology care. Individuals who identify as non-Hispanic Black or as Hispanic may be more likely to live in higher poverty neighborhoods and experience factors that impact access to outpatient CHD specialty care which may contribute to higher inpatient and ED utilization and adverse outcomes. Those living in higher poverty neighborhoods may experience additional challenges accessing congenital cardiology care due to school/work environment, availability and quality of health care services, insurance coverage, health literacy, and community resource constraints.^20, 21^ Our study also appears to demonstrate disparities by age and race and ethnicity in individuals who have had any type of healthcare encounter. It has been reported that children with CHD who identify as Black or Hispanic are less likely to have cardiology follow-up and remain in cardiology care,^13, 42^ which may be related to a variety of issues impacting access to care such as health insurance, transportation, language barriers, implicit bias in the healthcare system, and other social determinants.

Recent recommendations with policy implications for improving healthcare delivery for individuals with CHD recognize the role of individuals’ socioeconomic factors, in addition to healthcare delivery and workforce improvements.^43^ A framework focusing on access to care, affordability, and accessibility for all populations with CHD has been proposed as a vision for 2030, along with engaged leaders and identification of areas of improvement, training future workforce, and addressing barriers to care.^43^ Our findings support the complex interplay between race and ethnicity and neighborhood poverty status in healthcare utilization and mortality among individuals with CHD. Addressing these would require solutions related to expanded accessibility for lifelong CHD care, improved Medicaid funding or universal healthcare, and understanding issues related to patients, as recommended by Chowdhury et al. (2021). Mainly, our study supports efforts highlighted by Chowdhury et al. for addressing and assessing health care disparities including consistency in care and resources available for minority populations.

Our analysis is strengthened by the large study size spanning multiple geographic regions within the U.S. and diverse health administration data sources and vital records. We included both pediatric and adult healthcare systems and a broad age spectrum. Similar to other studies of CHD using administrative data, our dataset may contain individuals without CHD and may miss individuals with CHD, although in contrast to other studies using administrative data, we constrained our CHD codes to those with higher positive predictive value. We could examine CHD anatomic grouping as a co-variable in understanding racial and ethnic disparities by neighborhood poverty status for different types of healthcare utilization and mortality. However, there are some key limitations. Overall, 12% of individuals were excluded who met the diagnostic criterion, but who did not have information on their race and ethnicity. We conducted separate analyses by including those individuals with “unknown” race and ethnicity (**Table S1**), and while the majority of findings did not change remarkably from the current analysis, for individuals with “unknown” race residing in either high or low poverty neighborhoods, the odds of having a hospitalization or having an emergency room visit was less compared to white individuals (see **Table S2**). However, heterogeneity of measurement, aggregation of multiple racial groups into “other” category, and missing data issues on race and ethnicity pose challenges when ascertaining and interpreting our findings on healthcare utilization disparities in select minority populations.^44^ Limited availability of racial and ethnic data not only affects the current analysis, but also is necessary to the measurement, identification, understanding and ultimately, the elimination of disparities in health as well as to the improvement of the quality of healthcare in a standardized way for all individuals.^45, 46, 47, 48^ We used eHR data which misses individuals who did not seek medical care. We did not have data on type of outpatient visit, e.g., receipt of congenital cardiology care, limiting our ability to examine variability in the type of outpatient care individuals received. Mortality among individuals with CHD was based on state-specific vital records at each site, rather than NDI, and was limited to the three-year surveillance period. Our reliance on state death certificates may have underestimated mortality over the surveillance period, especially if some deaths occurred outside the catchment regions. Additionally, a longer longitudinal analysis of healthcare utilization and survival in this population would enhance our understanding of healthcare utilization and its effect on the risk of mortality. In addition, there could be multiple reasons for hospitalizations among individuals with CHD, and these reasons can be age-specific.^39^ Four age groups were examined in the current study, including children (1-10 years) adolescents (11-18 years) and two groups of adults: 19-44 years and 45-64 years. While these groupings allow us to examine age-related effects, more granular analysis of age by race and ethnicity would be possible with larger sample sizes, specifically among older aged categories.

In conclusion, our study showed that healthcare utilization was associated with race and ethnicity among individuals with CHD in both low and high poverty neighborhoods, and with mortality in high poverty neighborhoods. Assessing community-level social determinants of health, along with access to healthcare, may help close the gaps in racial and ethnic inequities in healthcare utilization and mortality among individuals with CHD.

## Data Availability

Contact Jill Glidewell at Centers for Disease Control and Prevention for data availability

## Acknowledgements

The authors would like to thank all collaborators from each participating site and CDC: Trenton Hoffman, Carol Hogue, Daphne Hsu, Aida Soim, Alissa Van Zutphen, Kirstin Sommerhalter, Ali Zaidi, Sergey Krikov, Matthew Reeder, Kevin Whitehead, Grace Ryan, Karen Chiswell, Tracy Spears, Lauren Sarno, Timothy Hoffman, Karl Welke, Michael Walsh, Tiffany Colarusso, Karrie Downing, Bobby Lyles. The authors would also like to thank the Intermountain Healthcare’s Adult Congenital Disease Program and all institutions that contributed data to the project.

**Appendix A.** ICD-9-CM Codes for Anatomic Complexity of Congenital Heart Defects, 63 codes.

| Classification | ICD-9-CM Code | ICD-9-CM Description |
| --- | --- | --- |
| **Severe**<br>**16 codes**<br>(contains at least one severe code) | 745.0 | Common truncus |
|  | 745.1 | Transposition of great vessels |
|  | 745.10 | Transposition of great vessels: Complete transpos. great vessels |
|  | 745.11 | Transposition of great vessels: Double outlet right ventricle |
|  | 745.12 | Transposition of great vessels: Corrected transpos. great vessels |
|  | 745.19 | Transposition of great vessels: Other |
|  | 745.2 | Tetralogy of Fallot |
|  | 745.3 | Common ventricle |
|  | 745.6 | Endocardial cushion defects |
|  | 745.60 | Endocardial cushion defects: Endocard cushion, unsp. type |
|  | 745.69 | Endocardial cushion defects: Other |
|  | 746.01 | Anomalies of pulmonary valve: Atresia, congenital |
|  | 746.1 | Tricuspid atresia and stenosis, congenital |
|  | 746.7 | Hypoplastic left heart syndrome |
|  | 747.11 | Coarctation of aorta: Interruption of aortic arch |
|  | 747.41 | Anomalies great veins: Tot anomalous pulm. venous connection |
| **Shunt**<br>**7 codes**<br>(contains at least one shunt code and no severe or valve codes) | 745.4 | Ventricular septal defect |
|  | 745.5** | Ostium secundum type atrial septal defect** |
|  | 745.61 | Endocardial cushion defects: Ostium primum defect |
|  | 745.8 | Bulbus cordis anomalies & anomalies of card septal closure: Other |
|  | 745.9 | Unspecified defect of septal closure |
|  | 747.0 | Patent ductus arteriosus |
|  | 747.42 | Anomalies great veins: Partial anomal. Pulm. venous connection |
| **Valve**<br>**17 codes**<br>(contains at least one valve code, and no severe or shunt codes) | 746.0 | Anomalies of pulmonary valve |
|  | 746.00 | Anomalies of pulm valve: Pulm valve anomaly, unsp. |
|  | 746.02 | Anomalies of pulmonary valve: Stenosis, congenital |
|  | 746.09 | Anomalies of pulmonary valve: Other |
|  | 746.2 | Ebstein's anomaly |
|  | 746.3 | Congenital stenosis of aortic valve |
|  | 746.4 | Congenital insufficiency of aortic valve |
|  | 746.5 | Congenital mitral stenosis |
|  | 746.6 | Congenital mitral insufficiency |
|  | 746.81 | Other specified anomalies of heart: Subaortic stenosis |
|  | 746.83 | Other specified anomalies of heart: Infundibular pulmonic stenosis |
|  | 747.1 | Coarctation of aorta |
|  | 747.10 | Coarctation of aorta: Coarctation of aorta (preductal) (postductal) |
|  | 747.22 | Other anomalies of aorta: Atresia and stenosis of aorta |
|  | 747.3 | Anomalies of pulmonary artery |
|  | 747.31 | Anomalies of pulmonary artery: Pulm. artery coarctation & atresia |
|  | 747.39 | Anomalies pulmonary artery: Anomal. pulm. artery & pulm. Circ. |
| **Shunt and Valve** (contains at least one shunt code and at least one valve code, and no severe codes) |  |  |
| **Other**<br>**(23 codes)**<br>(contains one or more codes listed in group, without severe, shunt or valve codes)<br>Excluded from current analysis | 648.5 | Other current conditions in mother classifiable elsewhere, but complicating preg., childbirth, or puerperium: Congen. cardio dis. |
|  | 648.50 | Other current conditions in mother classifiable elsewhere, but complicating preg., childbirth, or puerperium: Congen. cardiovasc. dis.: unspecified as to episode of care or not applicable |
|  | 648.51 | Other current conditions in mother classifiable elsewhere, but complicating preg., childbirth, or puerperium: Congen. cardiovasc. dis.: delivered, w/ or w/o mention of antepartum condition |
|  | 648.52 | Other current conditions in mother classifiable elsewhere, but complicating preg., childbirth, or puerperium: Congen. cardiovasc. dis.: delivered, w/mention of postpartum complication |
|  | 648.53 | Other current conditions in mother classifiable elsewhere, but complicating preg., childbirth, or puerperium: Congen. cardiovasc. dis.: antepartum condition or complication |
|  | 648.54 | Other current conds. in mom classifiable elsewhere, but complic. preg., childbirth, or puerperium: congen. cardio. dis.: PP conds. |
|  | 745.7 | Cor biloculare |
|  | 746.8 | Other spec. anomalies of heart |
|  | 746.82 | Other spec. anomalies of heart: Cor triatriatum |
|  | 746.84 | Other spec. anomalies of heart: Obstructive anomalies, NEC |
|  | 746.85 | Other spec. anomalies of heart: Coronary artery anomaly |
|  | 746.87 | Other spec. anomalies of heart: Malposition of heart & cardi apex |
|  | 746.89 | Other specified anomalies of heart: Other |
|  | 746.9 | Unspecified anomaly of heart |
|  | 747.2 | Other anomalies of aorta |
|  | 747.20 | Other anomalies of aorta: Anomaly of aorta, unspecified |
|  | 747.21 | Other anomalies of aorta: Anomalies of aortic arch |
|  | 747.29 | Other anomalies of aorta: Other |
|  | 747.4 | Anomalies of great veins |
|  | 747.40 | Anomalies of great veins: Anomaly of great veins, unspecified |
|  | 747.49 | Anomalies of great veins: Other anomalies of great veins |
|  | 747.9 | Unspecified anomaly of circulatory system |
|  | V13.65 | Congenital (corrected) malformations: Personal hx of (corrected) congenital malformations of heart and circulatory system |

\* adopted from Lifespan CHD surveillance project (Glidewell JM, Farr SL, Book WM, Botto L, Li JS, Soim AS, Downing KF, Riehle-Colarusso T, D'Ottavio AA, Feldkamp ML, Khanna AD, Raskind-Hood CL, Sommerhalter KM, Crume TL. Individuals aged 1-64 years with documented congenital heart defects at healthcare encounters, five U.S. surveillance sites, 2011-2013. *Am Heart J.* 2021 Aug;238:100-108).
\*\* 745.5 in isolation or with 'Other' codes have been omitted from analyses due to lack specificity
**Notes.** **Severe** has at least one severe code, regardless of presence of shunt, valve, or shunt and valve codes. **Shunt and Valve** has shunt AND valve codes; **Shunt** has at least one shunt code, and no valve or severe codes. **Valve** has at least one valve code, and no shunt or severe codes. **Other** has one+ codes in this category, but this category was omitted from analyses due to non-specificity. **745.5** has either one code, 745.5, or code 745.5 in addition to having only codes from the 'Other' category (see Table above); this category was also omitted from analyses due to non-specificity.

---

**Table S1.** Characteristics of individuals with congenital heart defects, total and by racial and ethnic group.

|  | Total<br>N=35888 | Non-Hispanic<br>White<br>n=18173<br>(50.6%) | Non-Hispanic<br>Black<br>n=6991<br>(19.5%) | Hispanic<br>n=5365<br>(15.0%) | Other<sup>‡</sup><br>n=1013<br>(2.8%) | Unknown<br>n=4346<br>(12.1%) | P-value<sup>§</sup> |
| --- | --- | --- | --- | --- | --- | --- | --- |
| Characteristics | n (%) | n (%)<sup>*</sup> | n (%)<sup>*</sup> | n (%)<sup>*</sup> | n (%)<sup>*</sup> | n (%)<sup>*</sup> |  |
| Site |  |  |  |  |  |  |  |
| Georgia | 8165 (22.8) | 2854 (15.7) | 2415 (34.5) | 522 (9.7) | 279 (27.5) | 2095 (48.2) | <0.0001 |
| North Carolina | 12601 (35.1) | 8033 (44.2) | 2454 (35.1) | 981 (18.3) | 362 (35.7) | 771 (17.7) |  |
| New York | 10778 (30.0) | 4247 (23.4) | 2091 (29.9) | 3534 (65.9) | 368 (36.3) | 538 (12.4) |  |
| Utah | 4344 (12.1) | 3039 (16.7) | 31 (0.5) | 328 (6.1) | <10 (--) | 942 (21.7) |  |
| Age<sup>†</sup> (years) |  |  |  |  |  |  |  |
| 1-10 | 14804 (41.2) | 5815 (32.0) | 3585 (51.3) | 2765 (51.5) | 535 (52.8) | 2104 (48.4) | <0.0001 |
| 11-18 | 7497 (20.9) | 3842 (21.2) | 1311 (18.7) | 943 (17.6) | 155 (15.3) | 1246 (28.7) |  |
| 19-44 | 8353 (23.3) | 5076 (27.9) | 1333 (19.1) | 1096 (20.4) | 200 (19.8) | 648 (14.9) |  |
| 45-64 | 5234 (14.6) | 3440 (18.9) | 762 (10.9) | 561 (10.5) | 123 (12.1) | 348 (8.0) |  |
| Sex |  |  |  |  |  |  |  |
| Female | 17432 (48.6) | 8295 (45.6) | 3718 (53.2) | 2834 (52.8) | 529 (52.2) | 2056 (47.3) | <0.0001 |
| Male | 18456 (51.4) | 9878 (54.4) | 3273 (46.8) | 2531 (47.2) | 484 (47.8) | 2290 (52.7) |  |
| CHD Anatomic Severity |  |  |  |  |  |  |  |
| Severe | 8693 (24.2) | 4488 (24.7) | 1829 (26.2) | 1230 (22.9) | 268 (26.5) | 878 (20.2) | <0.0001 |
| Shunt | 10609 (29.6) | 4163 (22.9) | 2452 (35.1) | 2076 (38.7) | 376 (37.1) | 1542 (35.5) |  |
| Valve | 13664 (38.1) | 8145 (44.8) | 2052 (29.3) | 1601 (29.8) | 261 (25.8) | 1605 (36.9) |  |
| Shunt and Valve | 2922 (8.1) | 1377 (7.6) | 658 (9.4) | 458 (8.6) | 108 (10.6) | 321 (7.4) |  |
| Health Insurance |  |  |  |  |  |  |  |
| Any public | 17830 (49.7) | 6157 (33.9) | 4987 (71.3) | 4399 (82.0) | 528 (52.1) | 1759 (40.5) | <0.0001 |
| Private | 14791 (41.2) | 10063 (55.4) | 1440 (20.6) | 700 (13.0) | 358 (35.3) | 2230 (51.3) |  |
| None | 370 (1.0) | 148 (0.8) | 87 (1.3) | 67 (1.3) | <10 (--) | 60 (1.4) |  |
| Unknown | 2897 (8.1) | 1805 (9.9) | 477 (6.8) | 199 (3.7) | 119 (11.7) | 297 (6.8) |  |
| Neighborhood median income (USD) |  |  |  |  |  |  |  |
| <\$40,000 | 10922 (30.4) | 3295 (18.1) | 3313 (47.4) | 3039 (56.7) | 356 (35.1) | 919 (21.2) | <0.0001 |
| \$40,000-\$75,000 | 19471 (54.3) | 11142 (61.3) | 3398 (48.6) | 2035 (37.9) | 456 (45.0) | 2440 (56.1) | |
| >\$75,000 | 5495 (15.3) | 3736 (20.6) | 280 (4.0) | 291 (5.4) | 201 (19.9) | 987 (22.7) | |
| Neighborhood poverty status |  |  |  |  |  |  |  |
| Low poverty neighborhood<br>(<25% households below FPL) | 24430 (68.1) | 14684 (80.8) | 3587 (51.3) | 2328 (43.4) | 635 (62.7) | 3196 (73.5) | <0.0001 |
| High poverty neighborhood<br>(≥25% households below FPL) | 11458 (31.9) | 3489 (19.2) | 3404 (48.7) | 3037 (56.6) | 378 (37.3) | 1150 (26.5) |  |
| Healthcare utilization |  |  |  |  |  |  |  |
| Outpatient visits | 33381 (93.0) | 16730 (92.1) | 6532 (93.4) | 5175 (96.5) | 915 (90.3) | 4029 (92.7) | <0.0001 |
| Hospitalizations | 14431 (40.2) | 7318 (40.3) | 3099 (44.3) | 2599 (48.4) | 507 (50.1) | 908 (20.9) | <0.0001 |
| Emergency Department visits | 11313 (31.5) | 4827 (26.6) | 2453 (35.1) | 3226 (60.1) | 272 (26.9) | 535 (12.3) | <0.0001 |
| Mortality |  |  |  |  |  |  |  |
| Yes | 399 (1.1) | 197 (1.1) | 115 (1.6) | 50 (0.9) | 12 (1.2) | 25 (0.6) | <0.0001 |

**Acronyms:** CHD = congenital heart defects; FPL = Federal Poverty Level; USD = United States dollar currency.
\* Column percentages reported in cells.
† Estimates for 1-10-year-olds based on three sites: five counties in metropolitan-Atlanta, GA; 11 counties in NY state; and statewide in NC; estimates for 11-18, 19-44, and 45-64-year-olds based on four sites: five counties in metropolitan-Atlanta, GA; 11 counties in NY; and statewide in NC and UT.
‡ Other racial and ethnic group includes Asian, American Indian/Native American, Native Hawaiian or Other Pacific Islander, and multi-racial.
§ Comparing racial and ethnic groups only; analysis does not include “Total” column.
|| Insurance categorization: public defined as Medicaid or Medicare; private defined as private, other government, or other insurance; none defined as self-pay/uninsured; and unknown defined as unavailable, unknown, or no insurance indicated.

**Table S2.**
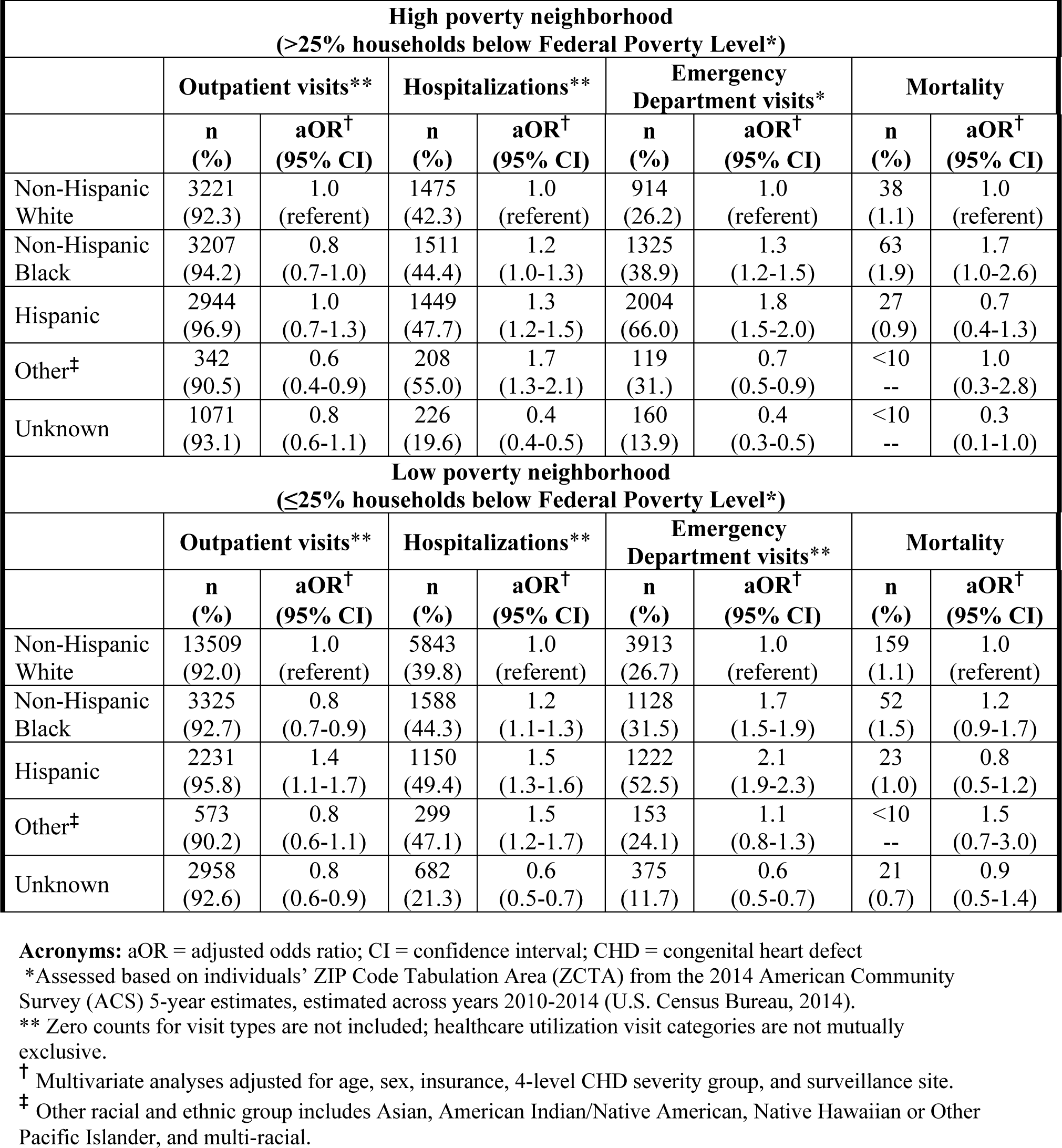
Associations between race and ethnicity and healthcare utilization and mortality, by neighborhood poverty status among individuals with congenital heart defects.

